# It’s complicated…: Exploring the missed opportunities and reasons for non-performance of assisted vaginal births in Kenya

**DOI:** 10.1101/2022.12.21.22283818

**Authors:** Fiona M Dickinson, Helen Allott, Paul Nyongesa, Martin Eyinde, Onesmus M Muchemi, Stephen W Karangau, Evans Ogoti, Nassir A Shaban, Pamela Godia, Lucy Nyaga, Charles A Ameh

## Abstract

Unnecessary Caesarean Section (CS) can have adverse effects on women and their newborn. Assisted vaginal birth/delivery (AVB/AVD) using a suction device or obstetric forceps is a potential alternative when delays or complications occur in the second stage of labour. Unlike CS, AVB using a suction device does not require regional or general anaesthesia, can often be performed by midwives, and does not scar the uterus, lowering the risk of maternal mortality and morbidity, in this and subsequent pregnancies. This study examined the justification for, and outcomes of second stage CS (SSCS) and reasons for low levels of use of AVB, in Kenya.

Using a mixed methods study design, we reviewed case-notes from women having AVB and second-stage CS births, and conducted key informant interviews with healthcare providers, from 8 purposively selected hospitals in Kenya. Randomly selected SSCS and all AVB case-notes were reviewed by a panel of four experienced obstetricians, and appropriateness of the procedure assessed. Semi-structured interviews were conducted and analysed using a thematic approach.

Review of 67 SSCS case-notes showed 10% might have been conducted as AVBs, with a further 58% unable to be classified due to inadequate/inconsistent record keeping or excessive delay following initial CS decision. Outcomes following SSCS showed perinatal mortality rate of 89.6/1,000 births, with 11% of infants and 9% of mothers experiencing complications. Non-referred cases of AVB showed good outcomes. Twenty interviews were conducted with obstetricians, medical officers and midwives. The findings explored the experience and confidence of healthcare providers in performing AVBs, and adequacy of the training they received. Key reasons for non-performance included lack of functioning equipment, lack of trained staff or their rotation to other departments.

Reasons for non-performance of AVB were complex and often multiple. Any solutions to these problems will need to address various local, regional and national issues.

## Background

Key interventions to shorten the second stage of labour i.e. after full dilation of the cervix, include assisted vaginal birth (AVB) and caesarean section (CS), both of which are important aspects of emergency obstetric care [1]. They may be indicated for maternal fatigue, maternal medical indications, suspected fetal hypoxia or prolonged duration of the second stage. Failure to intervene, when necessary, could be associated with poor outcomes for both mother and baby [2], however, birth by CS has also been found to have a significant association with poor maternal outcomes, especially when performed in the second stage of labour [3].

In their systematic review and meta-analysis of CS in low and middle-income countries (LMIC) Sobhy et al [3], found a CS associated risk of maternal mortality of 10.9 per 1,000 procedures in sub-Saharan Africa compared to 0.3 per 1,000 procedures in LMIC in Europe and central Asia. Emergency CS in all LMIC were twice as likely to result in a maternal death compared to elective CS, and CS performed in the second stage of labour (SSCS), was 12 times more likely to end in maternal death compared to those done in first stage [3]. Other adverse maternal and neonatal outcomes were also significantly increased with SSCS, including admission to ITU, hysterectomy and perinatal death [3]). In addition to mortality and short-term morbidity, there are also longer-term risks associated with caesarean section, including uterine rupture in subsequent labour [4] and abnormal placentation [5]. Given these risks it is important that caesarean birth, especially SSCS, is restricted to those situations where there is no safe alternative means of delivery, and unnecessary CS are avoided.

Depending on the level of descent of the fetal head, in many cases delivery by means of AVB is an alternative to SSCS. A large cohort study from Uganda [6] demonstrated significantly fewer maternal complications after AVB than after SSCS (0.8% v’s 4.2%, *p*=0.003), whereas perinatal outcomes, including deaths overall, were comparable for the two groups. During the decision to delivery interval (DDI), significantly more fetal deaths occurred in the SSCS group than AVB group (0.9% v’s 4.4%, *p*=0.003) [6]. This may, at least in part, have been due to the higher proportion of women having a DDI of >60 minutes in the SSCS group compared to the AVB group (29.3% v’s 81.9%, p=<0.001). An earlier study in UK replicated many of these findings, with higher levels of maternal morbidity following SSCS compared to AVB and more neonatal admissions to the special care baby unit, although there was less neonatal trauma reported with SSCS [7]. A retrospective study of hospital records in Nigeria, reported significant reductions in perinatal deaths (*p*=0.04) and fetal deaths during the decision to delivery interval (*p*=0.029) relating to AVB compared to SSCS, although other newborn outcomes showed no statistically significant difference [8].

It is surprising that, given the evidence that in LMIC, SSCS carries greater maternal risks and at best, equivalent fetal risks, relatively few AVBs are conducted in many low-resource settings, especially in sub-Saharan Africa [9]. The ability to perform AVB is a basic emergency obstetric care function that should be provided in all healthcare facilities providing intrapartum care. However, in one large multi-country study of LMIC, in sub-Saharan Africa AVB was found to have taken place within the previous three months, in only 53% of 1728 hospitals and 6% of nearly 10,000 health centres surveyed [9]. Overall institutional rates of AVB were around 1% and it was clear that in a great many hospitals and health centres AVB was not being practiced at all. Reasons for a lack of AVB included a lack of available or trained staff, a lack of available functional equipment and policies restricting authorisation to perform the procedure.

Since 2019, the Liverpool School of Tropical Medicine (LSTM) has been providing emergency obstetric and newborn care (EmONC) training, including AVB, to healthcare providers in 25 healthcare facilities across five counties in Kenya. Despite having trained 1193 members of staff, analysis of data routinely collected from supported healthcare facilities, revealed that the rates of AVB remained very low. The data showed that in the eight highest volume facilities, during the period June to November 2020, 12,161 births occurred. Of these, 25.6% (n=3114) of births were conducted by CS whilst only 0.31% (n=38) were conducted by AVB, with three of the eight facilities not conducting any AVBs.

The objectives of this study were to explore the missed opportunities and outcomes of SSCS and understand clinician’s perceptions and practice regarding the use or non-use of AVB, in selected high-volume facilities, in Kenya.

## Methods

This study used a mixed methods approach, combining a cross-sectional, retrospective review of randomly selected case-notes from women who had undergone either SSCS or AVB, with key informant interviews with healthcare providers involved in the provision of intrapartum care. Ethical approval was sought and obtained from the LSTM Research Ethics Committee (ref: 21-041)and the Moi University Institutional Research Ethics Committee (ref: IREC/2021/115). Written informed consent was obtained from all study participants.

### Study setting

Case notes and interviewees were drawn from hospitals which were part of a Foreign, Commonwealth & Development Office funded, Maternal and Newborn Health Programme implemented in five of the 47 counties across Kenya. As the most frequently cited reason for non-performance of AVB was a lack of clients needing the procedure, eight hospitals were selected from the 25 healthcare facilities in the programme, that had the highest number of births (an average of more than 130 births per month). All included healthcare facilities were either county level or sub-county hospitals, with seven operating at a comprehensive Emergency Obstetric and Newborn Care (CEmOC) level (able to offer CS and AVB) and one at a basic EmONC (BEmOC) level, offering AVB but not CS. The lower-level health facility because it had a high volume of births per month and might be more likely to perform AVB because it did not provide CS.

### Sample size and data collection

#### Case-note review

The study period of June to November 2020, was chosen to avoid the restrictions surrounding the initial onset of the Coronavirus pandemic and a healthcare providers strike which occurred in December 2020. CS case-notes were selected using a systematic random sampling technique, using either the Kenya Health Information System, where possible, or the hospital maternity registers, as a sampling frame.

A sample size of 300 women who had a SSCS was initially planned, to ensure that the margin of error (MOE) in estimation of the proportion of women for whom an AVB could have been done would not exceed 5.7%. If 10% (or 5%) of women who had a SSCS had an adverse outcome, estimation of this proportion would have a MoE of 3.4% (or 2.5%).

Initial sample size calculations were based on stratified sampling of elective, first stage and SSCS case-notes but due to difficulties in identifying appropriate cases and locating the relevant notes in hospitals, a simple random sample of all emergency CS (first and second stage CS) notes was taken. A target of 90 emergency CS cases per hospital was set for the seven CEmOC hospitals, giving an overall sample of 630 emergency CS cases.

Once case-notes were identified and located, they were copied by hospital staff and sent to a central office in Nairobi. Here they were scanned, given a unique study ID number, and personal identifiable information was electronically redacted. In addition, due to the small number of procedures performed, all AVB cases for the study period, were included in the data collection.

#### Key informant interviews

For the key informant interviews (KII), purposive sampling was employed to identify nurse-midwives (also sometimes referred to as nurses), medical officers and obstetricians who provided clinical care in each of the eight target hospitals. A range of clinicians who were actively involved in the provision of care to women giving birth were identified, in addition to those with managerial responsibility, such as medical superintendents and midwives in-charge of labour wards. The aim was to identify staff who might be expected to either perform AVB or in a position to influence its use by others.

Using a semi-structured topic guide, the interviews were conducted remotely, due to the Covid-19 pandemic and logistical challenges, via the participants online platform of choice. Potential participants were identified by members of the programme team working with the facilities. These were screened by members of the study team and purposively selected to ensure a spread of cadres and facility representation. Participants were sent an invitation to take part in the study along with the participant information sheet and a consent form. They were given the opportunity to ask any questions of the research team before signing and returning the consent form. A date and time convenient to the participant was then arranged for the interview.

### Analysis

Data relevant to the method of birth was extracted from the CS and AVB case-notes using a pro forma and reviewed by a panel of four experienced obstetricians, three from Kenya and one from the UK but with experience of working in LMIC including Kenya. Each case was independently reviewed by two members of the panel and a conclusion drawn as to the potential appropriateness of the procedure. Where any discrepancies were identified between panellists, they were discussed by the group as a whole and a consensus formed. Appropriateness of CS, based on the available information, was classified as:

1. **CS** was considered to have been necessary and **appropriate**
2. **Unclear** due to lack of information in the notes
3. **Unclear** due to delay in performing the procedure
4. **AVB** should have been attempted

Data were also extracted from the notes on maternal and neonatal outcomes following both SSCS and AVB. These were analysed as simple descriptive statistics.

The KIIs were recorded and transcribed, and framework analysis undertaken [10]. This process involved familiarisation with the data through reading transcripts and listening to recordings, followed by thematic analysis to develop codes which are systematically applied to the data. The data were then summarised into charts, facilitating mapping and interpretation of the data. This process was carried out using qualitative data analysis software (NVivo, version 12).

## Results

From a total of 826 randomly selected CS case-notes retrieved, 223 (27.0%) were elective CS (before onset of labour), 507 (61.3%) were carried out in the first stage of labour, 67 were identified as SSCS (8.1%) and in a further 29 cases (3.3%) it was not possible to determine from the notes, the stage of labour in which the CS was performed. In addition, 6 sets of AVB case notes were identified and retrieved.

### Missed opportunities and outcomes

Following review of the available data for the 67 SSCS, in 22 cases (33%) SSCS was considered appropriate (**Figure 1**). These included cases of fetal arm prolapse and transverse lie, as well as instances where the head was still more than two fifths palpable abdominally. In the two ‘unclear’ categories, there were 21 cases (31%) where a judgement could not be made by the expert panel as to the appropriateness of SSCS, due to lack of information or contradictions in the medical records. There were also 17 cases (25%) where there was a significant delay between making the decision to perform CS and the procedure being performed. In two of these cases, there was a 5-hour delay from decision to delivery, with no reassessment recorded. It was felt that in the ‘Unclear due to delay in performing the procedure’ cases, further fetal descent might have occurred making AVB possible, thus expediting delivery and eliminating the need for SSCS.

**Figure 1.**
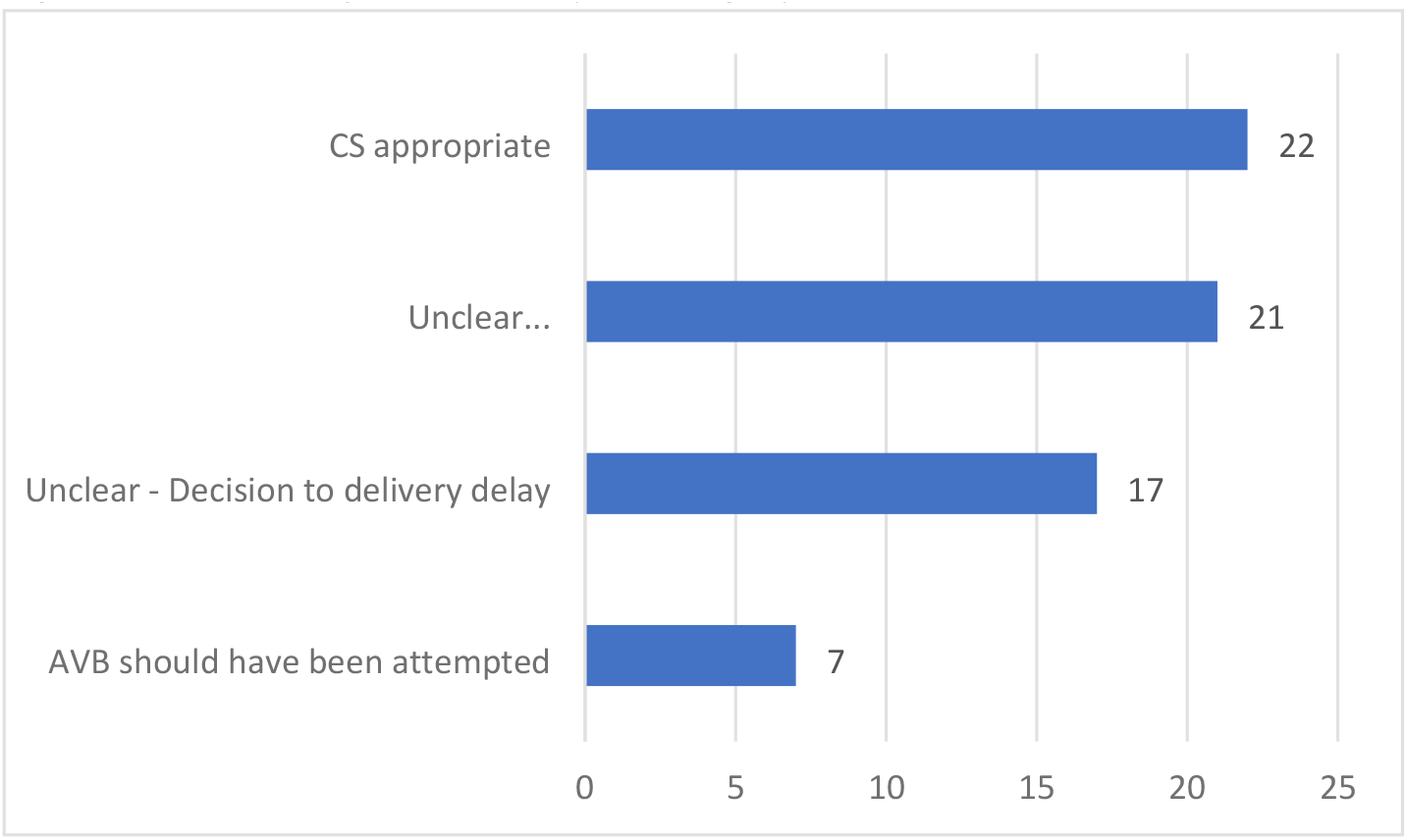
Number of SSCS cases per category

In the final category, there were 7 cases (10%) where reviewers felt that based on the case-notes, AVB should definitely have been attempted. Of these, 6 (86%) were ‘referred in’ from other facilities and 3 (43%) had a decision to delivery interval (time from making a decision to perform CS to the actual birth) of 2 hours or more. Three of the seven cases also had a total second stage (time from full dilation to birth) of at least 4 hours with 2 cases of at least 7 hours.

#### Maternal and perinatal outcomes following SSCS

Due to the small number of SSCS case-notes included in the records retrieved, statistical analysis was limited. However, analysis of the outcomes for the 67 SSCS cases revealed that there were 6 cases (9%) of maternal complications, including 5 (8%) of postpartum haemorrhage (PPH) but no maternal deaths (**Table 1**). Perinatal outcomes among the SSCS showed 2 (3%) neonatal deaths, 4 (6%) stillbirths which would equate to a perinatal mortality rate of 89.6 deaths per 1,000 births. A further 7 (11%) infants had complications needing treatment and/or admission to the neonatal unit (e.g. for convulsions or asphyxia). Overall birthweights ranged from 1705g to 4470g but did not seem to be associated with adverse outcomes for either mother or baby. Apgar scores for infants born alive and where data was available, was similar across all five groups.

**Table 1.** Maternal and perinatal outcomes

| Mode of birth/<br>assessment of<br>appropriateness of<br>SSCS (n) | Cases of<br>maternal<br>complications | Cases of<br>Stillbirth | Cases of<br>neonatal<br>death | Cases of<br>neonatal<br>complications | Mean birth<br>weight:<br>g (SD) | Mean Apgar<br>score @ 5<br>min:<br>score (SD) | Mean<br>decision to<br>delivery<br>interval:<br>hours (SD) |
| --- | --- | --- | --- | --- | --- | --- | --- |
| SSCS/AVB should have<br>been attempted (7) | 1 | 1 | 0 | 1 | 3244 (362) | 9 (2.5) | 1.9 (1.6) |
| SSCS/Unclear –<br>Decision to delivery<br>delay (17) | 2 | 2 | 0 | 1 | 3639 (434) | 8.2 (1.8) | 2.1 (0.9) |
| SSCS/Unclear -<br>inadequate/<br>contradictory<br>information (21) | 1 | 0 | 1 | 3 | 3113 (473) | 7.6 (2.5) | 1.3 (0.8) |
| SSCS/CS appropriate<br>(22) | 2 | 1 | 1 | 2 | 3083 (554) | 8.5 (2.2) | 2.2 (2.1) |
| AVB/not applicable (6) | 1 | 0 | 0 | 2 | 3048 (385.6) | 8.3 (2) | Not<br>documented |

The most common reason given for SSCS was failure to progress or delay in second stage (n=36, 54.6%), however, 6 also reported fetal distress or non-reassuring fetal signs. Meconium was also listed as a reason for SSCS in one case although, despite the decision to delivery interval being 2 hours, the baby had Apgar scores of 9 at 1 minute and 10 at 5 minutes. Other reasons given for SSCS included issues with presentation such as arm prolapse, transverse lie and breech, and previous CS including one with a known stillbirth.

#### AVB cases

Six cases of AVB were reviewed, five of which occurred in sub-county hospitals and only one in a county referral hospital. There were no perinatal or maternal deaths with most procedures being performed for either ‘delay in second stage of labour’ or ‘poor maternal effort’. Four of the cases had good outcomes for the mothers and babies, with Apgar scores at 5 minutes of 9 or 10. The two remaining cases, however, were both referrals from approximately 30 minutes away, who were referred for delay in second stage of labour. One of the referred mothers experienced complications with bleeding postpartum which was managed by vaginal packing. Both infants were approximately 2.5kg in weight and had birth asphyxia with poor Apgar scores (5 and 7 at 5 minutes). One of the asphyxiated infants required admission to the Newborn Baby Unit for 5 days.

### Clinician’s perceptions and practice of AVB

Key informant interviews were conducted with 20 healthcare providers from eight high volume hospitals included in the study. Interviewees included nine nurses-midwives (NMW), of whom four were in-charge of labour ward or the maternity department; five medical officers (MO), one of whom was a superintendent; and six consultant obstetricians (OB). The interviews were carried out using Zoom and lasted on average 39 minutes (SD=10.45).

The interviews explored the views of clinicians and managers regarding the use of AVB and reasons why it was not used. Where possible, quotes have been provided to illustrate the findings of the study, but this was not always possible, due to the nature of the information, so as not to compromise the anonymity of the respondents. Where quotes are given, the interview number, cadre, and level of health facility (SCH – Sub County Hospital; CRH – County Referral Hospital), are also given to provide context.

#### Views on AVB

Most of the respondents acknowledged the benefits of AVB, with the avoidance of the attendant abdominal scar and potential complications associated with CS, particularly when performed in the second stage of labour. We asked respondents how they would feel about having an AVB themselves or for their wife/partner. Most said they would be accepting of the procedure providing that the necessary indications were met and that the operator was competent. They recognised the reduced risk of having an AVB rather than a CS.

> *“If I had the confidence, in whoever is attending to me, I would give a go ahead. Yeah because of instead of going for a caesarean, to nurse the scar when I can be done an AVD and deliver well*.*” (KII 5, NMW, SCH)*

Others had their doubts about the procedure, and one respondent stated that

> *“I wouldn’t let it get to that, I would have delivered her by caesarean before*.*” (KII 6, OB, SCH)*

Interviewees were also asked about their perceptions of the views of their local community regarding AVB. There were some differences of opinion, but most felt that the local community had no real understanding of about AVB and what it involved.

> *“Most of them are not learned, they don’t understand these things. Even if you get a consent, you tell them ‘mama I want to use this to help your baby come out, please co-operate’, they don’t understand*.*” (KII 1, MO, CRH)*

Some felt that the recipients would accept whatever was advised by the doctor or would end the pain of labour.

> *“Most of our ladies when you ask for their consent to do the assisted vaginal delivery, they just tell you, just do anything so that you can take out the baby. Because they are in so much pain at that time, so they will take anything that can be done so they can take out the baby*.*” (KII 17, MO, SCH)*

It was thought that some women would prefer AVB to CS, through fear of going to the operating theatre and a having an anaesthetic, and for others there was the concern of the potential limiting effect CS might have on the size of their family.

> *“The ones who want to give birth to fifteen children, they want to avoid caesarean section as much as possible because they know that is going to limit the number of children they are going to get. So, if you give them any other option apart from caesarean section they would gladly take it*.*” (KII 8, MO, CRH)*

A few interviewees felt that some women had a fear of the device and the potential damage it might cause to their baby or themselves, or had heard adverse reports from friends.

> *“Some clients they decline vacuum because of the, just misconceptions. I think maybe the stories in the community that once it’s done maybe the baby will come out with brain injuries*.*” (KII 3, NMW, CRH)*

There seemed to be little, in any, antenatal teaching for the women on different ways of giving birth, with explanations regarding AVB and its implications being provided when women were in advanced labour.

#### Experience and training

Of the 17 respondents who provided the information, 14 had conducted an AVB at some point in their career, with a few performing regularly, but others had not performed the procedure for two or more years.

Several of the interviewees had undergone EmONC training provided by LSTM, and a few had also done the LSTM Advanced Obstetric Surgical course or other AVB training. Both the standard and advanced LSTM courses cover theoretical and hands-on training on AVB, and overall, interviewees found the courses to be beneficial, particularly the practical aspects.

> *“Oh, I learnt something, it was helping me a lot. Because actually I did not know how to do it at first, but out of your training I managed to do two, two cases*.*” (KII 17, MO, SCH)*

However, despite most of the respondents having received AVB training previously, the need for additional training was commented on repeatedly. Some interviewees felt that they needed further training provided by external organisations such as LSTM or other NGOs, whilst others thought that on-the-job training would be preferable.

> *“I’ve done about three [AVB] since the training of [county], but I’ve done many from when I left medical school, but we were using the old equipment, we were not using the Kiwi. The Kiwi one I’ve used on three patients or so. I’m quite competent actually, personally I am okay, only a bit of refresher then everything will fall in place*.*” (KII 4, OB CRH)*

In-house training was reported in a few of the facilities, with accounts of continuing medical education (CME) sessions, where medical interns tended to present on theoretical aspects of a particular subject, with little or no opportunities to practice skills. Although these seemed to be open to all cadres, limited staffing levels meant that in practice staff were sometimes too busy to attend.

### Reasons for non-performance

During the routine facility assessments, the most common reason given for not performing AVB within a facility was a lack of patients for whom the intervention was indicated. However, this was rarely mentioned by interviewees, with the majority expressing that they thought more AVB should be performed. Several reasons were offered by respondents for not performing AVB, including a lack of functional equipment, insufficient staff, lack of knowledge, skill or confidence, lack of training and fear of complications.

#### Lack of equipment

The lack of operational equipment was a recurring theme, expressed by the interviewees as a reason for not performing AVB. One of the difficulties described in these settings, was that the preferred device for performing AVB, the Kiwi obstetric suction cup device, is designed to be single-use. However, due the cost of replacements and difficulties in procuring new devices, they are often sterilised and re-used.

> *“We sterilize using high level disinfection and we reuse it… I think it can do up to 12 patients”* (KII 11, NMW, CRH)

In some instances, though, this led to problems where the device was no longer functioning properly. On another occasion, an interviewee explained that problems with a dysfunctional device had led to a woman having to undergo a SSCS following a failed AVB, resulting in the loss of the baby.

When asked about the use of obstetric forceps, most respondents said that they either did not have them in their facility or that they were not used. One medical officer stated that they had used them once and a consultant admitted that they had never been trained in their use.

In two of the hospitals, the staff had requested replacement Kiwi devices, but they were not available, and they had been provided with Malmstrom vacuum extractors instead. Although these are reusable, they require two people to operate and were not favoured by the staff interviewed.

#### Staffing

The levels of staffing were discussed during the interviews, with almost all interviewees reporting inadequate levels. Interviewees commonly described 1-3 nurse-midwives on duty per shift with fewer at night than during the day. These nurse-midwives may have covered only the labour ward in the large County Referral Hospitals but in the smaller facilities, they were also expected to cover the antenatal and postnatal wards.

This shortage of staff was exacerbated in some instances by the rotation of experienced staff to different departments, either as part of the normal medical training process or because of industrial strike action. The frequency of rotation varied from weekly to 6 monthly resulting in a lack of AVB trained staff.

> *“The problem we have is high turnover of the MOs, so the ones we are having currently, I think they are not trained*.*”* (KII 3, NMW, CRH)

In one instance the rotation of non-specialist medical staff was reported as a reason for non-medically indicated CS, as the rotated doctor was more interested in practising their surgical skills.

The combination of low overall staffing levels and the rotation of trained staff was also reported as leading to a lack of support and on-the-job training opportunities for less experienced colleagues.

Instances were described where a shortage of MOs, lead to a lack of medical cover at times, resulting in the referral of women who developed complications in the second stage of labour.

> *“So at night it’s always nurses and if we find any difficulty we always refer*.*” (KII 2, NMW, SCH)*

This was also raised as an issue by receiving facilities, where due to delays caused by the women being transferred from one facility to another (up to 2 hours or more), AVB was not even attempted but the patient was taken straight to the operating theatre for caesarean section.

> *“This mother has travelled all the way more than 2 hours during second stage, I don’t wait, because I know, I just go direct and section this mother*.*” (KII 1, MO, CRH)*

Another reported effect of low staffing levels was the impact of the quality of patient care. Where a small number of nurse-midwives were caring for multiple women in labour, adequate monitoring of interventions such as augmentation of labour with oxytocin infusion was not possible. This resulted in the infusion being stopped, poor progress with failure to reach full dilatation or inadequate contractions and consequently the woman having a CS.

> *“You as a doctor might start the augmentation but then the nurses will stop it because they can’t just monitor. They will call you after four hours that the patient has not progressed… “(KII 4, OB, CRH)*

Where inadequate staffing was experienced, a few healthcare providers described the management of workloads by the juggling of patient care between cadres. Doctors were described as wanting an AVB to be done by the nurse-midwife, to avoid them having to take the patient to theatre, whilst the nurse-midwife wanted the woman to have a CS so that she was one less patient of labour ward.

#### Confidence

Varying levels of self-confidence were expressed by study respondents. Some were highly confident whilst others were less so. Levels of confidence were sometimes linked to training received and opportunities to practice. Respondents explained that their lack of confidence stemmed from a lack of any training or refresher training in performing AVB. They thought that with the necessary training they would be able to perform the procedure.

Confidence between colleagues was expressed as an issue by some respondents, with a few doctors conveying a lack of trust in some of the nurse-midwives’ ability to monitor the progress of labour whilst some of the nurse-midwives did not trust the doctors to perform an AVB when it was indicated.

A few of the respondents also articulated concerns they perceived colleagues had about potential adverse outcomes relating to the use of AVB, predominantly related to adverse outcomes for the mother and baby, particularly where the procedure was applied inappropriately.

> *“The thing we fear with the AVDs is the hematoma that sometimes is associated and possible dangers of having brain injuries as you do the AVDs and I think some of these fears are what many practitioners fear… If you follow the correct guidelines on who qualifies for AVDs nobody should fear*.*” (KII 18, MO, SCH)*

Two neonatal deaths were reported following AVB, possibly giving rise to some of the concerns, but it was difficult to determine whether these were because of inappropriate use of AVB itself or as a consequence of the labour complications which necessitated the AVB.

## Discussion

### Key results

Despite the provision of EmONC training, including AVB, in the project supported counties, routine programmatic data showed that the use of AVB remained very low. We reviewed a selection of SSCS and AVB case notes from the included healthcare facilities and found that 10% of SSCS births might have been carried out as AVB, potentially reducing the risk of complications and future morbidity for these women and infants following the procedures. We also found that, whilst many clinicians were in theory appreciative of the benefits of AVB, a complex array of factors impacted on their practice, including staffing, equipment and training.

### Strengths and limitations

This study obtained first-hand accounts from healthcare providers of different cadres, who worked clinically on labour wards, providing care for women giving birth. They included obstetricians, medical officers and nurse-midwives from eight of the highest volume healthcare facilities, in five counties, across different regions of Kenya. The data was collected in a secure manner, taking account of the ongoing Coronavirus pandemic, and using low-cost methods. We acknowledge that conducting the interviews remotely might have impacted the findings of this study, although the precise impact is difficult to determine. It is possible that remote interviews gave respondents an enhanced sense of anonymity, and enabled a greater degree of honesty, but conversely face-to-face interviews might have resulted in interviewers building a stronger rapport with respondents. Further research into the views of women who have given birth by AVB might also provide a broader, first-hand perspective on community level acceptability of AVB, but the scarcity of procedure might make recruitment of sufficient participants difficult.

Challenges in obtaining the notes for women who had SSCS resulted in a relatively small sample size for the case note review. In most healthcare facilities, a new set of hospital records was started for each pregnancy. Due to the way in which notes were filed it was not possible to reliably locate the SSCS cases, so it was necessary to retrieve a random selection of emergency CS and identify the SSCS cases at the review stage. Also, a major limiting factor in the interpretation of the SSCS and AVB cases was the quality of the record keeping. This varied considerably between facilities and in some cases had a notable impact on the reviewers’ ability to assess the need for, and outcomes of, the procedures. Examples of poor record keeping included incomplete or missing partographs, contradictions within the notes, or important information on labour progress not recorded.

### Interpretation of our results

#### Poor quality of records

Overall, the quality of medical record keeping was poor, with almost a third (32%, n=21) of the cases included in the sample not containing enough information to categorise the appropriateness of the decision for CS. Poor record keeping is not however limited to this study. Similar results have been found in other studies in LMICs including Landry et al [11] who reviewed records from CS performed in facilities in five low-income countries. They found in some sites 20% of records could not be located and key information was missing from files across all sites.

The lack of adequate filing systems and the incomplete or contradictory information that many of the sets of notes in this study contained, is likely to impact efforts to improve the quality of patient care, both directly as complete medical history is not available and through difficulty in conducting clinical audit and research [12]. Reasons for the poor quality of record keeping are likely to be various and a study of nurses’ knowledge, attitudes and barriers to quality record keeping in Burundi [13] found that barriers to good record keeping included lack of training in record keeping, excessive workload, lack of time, demotivation and poor support from administrative polices. Some of the facilities included in this study used a pre-formatted admission sheet which facilitated the systematic recording of important information. The inclusion of a completed partograph in the records was also variable, and unlikely where a woman had been referred from another facility, although no quantitative analysis has been done at this stage. A study carried out by Kleczka et al [14] in informal settlements in Nairobi, Kenya, found that the use of pre-formatted rubber stamp to print templates onto sheets of paper, significantly improved the quality of record keeping in three non-communicable disease clinics (hypertension, diabetes, and chronic respiratory diseases). The authors concluded that the intervention provided a low cost means of increasing the quality of medical documentation and supported improvement of care quality through standards-based audits.

#### Evidence and potential consequences of unnecessary SSCS

The number of cases in the group categorised as ‘AVB should have been attempted’ (n=7/67, 10%) suggests that there are women undergoing unnecessary SSCS. If the cases where it was not possible to determine the appropriate method of birth (due to lack of information or delays) are excluded, the proportion becomes even higher (n= 7/29, 24%). In this study, although there were complications following both SSCS and AVB, the small number of AVB performed made it impossible to compare the outcomes from the procedures statistically. Both of the AVB cases where complications occurred, may have been compromised due to the duration of referral following full dilatation and thus delays in actually performing the procedure. Furthermore, based on studies by Nolens et al [6] and Eze et al [8], if SSCS had been carried out rather than AVB, the further delay and complications from the procedure, may have resulted in worse outcomes.

There is no shortage of evidence around the increased risks and poor outcomes associated with unnecessary CS, particularly in LMIC [15],[3],[7],[16], both at the time of the procedure and in subsequent pregnancies. These risks are increased when the CS is carried out in the second stage of labour, after full dilatation of the cervix [8],[17].

There is little doubt about the need to avoid unnecessary CS in low resource settings, particularly SSCS, but there is also a need to ensure that those women who do need a CS, receive one in a timely manner. Arunda et al [14] highlight the demographic factors that impact on the likelihood of women receiving a CS in Kenya and Tanzania, including living in a rural or urban environment, household income and level of education. They advocate for the removal of financial incentives to conduct CS, reduction in unwanted, particularly adolescent pregnancies, and individualised decision making at a facility level, rather than blanket policies, to promote equitable access to necessary obstetric surgery. Factors such as patient referral, postpartum haemorrhage, the need for intensive care and general anaesthesia at CS were significantly associated with increased risk of maternal mortality following CS in Kenya [17].

Interventions to support decision making around CS have included mandatory second opinion [18], as well as the promotion of AVB where appropriate and raising awareness among women of the risks associated with unnecessary CS [20]. Dindi et al [18] advocate the prospective use of the Robson 10-group classification system for categorising CS, as a means of improving the quality of record keeping and underpinning efforts to improve care quality through audit, an intervention promoted by the WHO [21].

#### Complex reasons for non-performance of AVB

The qualitative findings from the study clearly demonstrated the complexity of the situations in which many doctors and nurse-midwives found themselves. Despite many clinicians expressing support for the performance of AVB, the data collected from the healthcare facilities did not demonstrate this in practice. The barriers to performing AVB described by the interviewees can be categorised at 3 levels, personal (confidence and practice), facility (equipment, on-job-training, leadership) and national (staffing and pre-registration training), illustrated in the conceptual diagram (Figure 2). These levels were not discrete however, with each level potentially having an impact on the other levels and some issues, such as training, being cross-cutting across all three levels.

**Figure 2.**
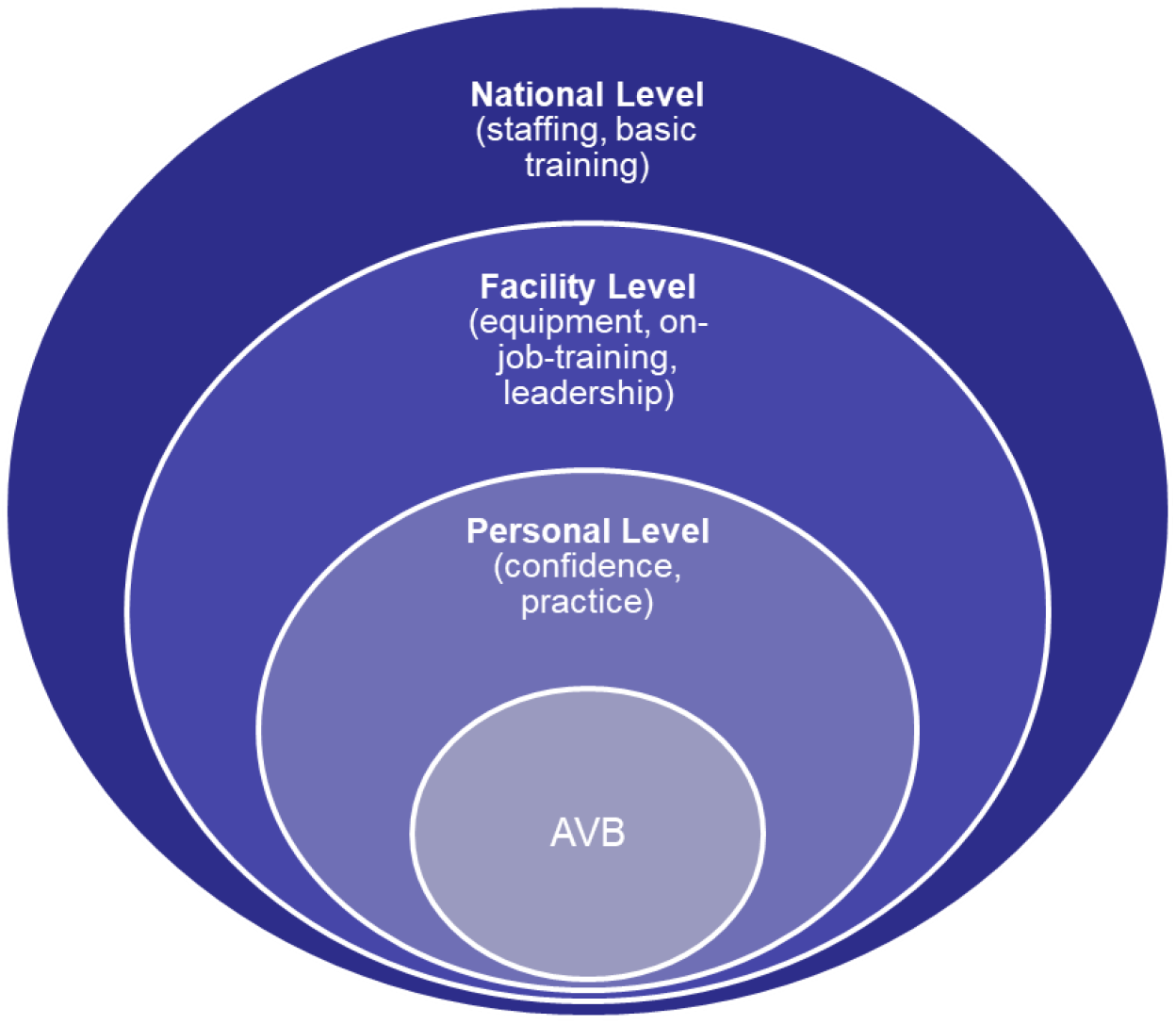
Conceptual diagram of reasons for non-performance of AVB

##### Equipment

The lack of functioning basic equipment was a barrier to the practice of AVB in some instances, and in line with the findings of research in other high, middle and low-income countries [9],[22],[23]. Staff generally expressed a preference for the Kiwi Omni-cup device as it was easy to use and only required a single individual to perform the procedure, but these devices are designed to be single-use. None of the interviewees used obstetric forceps and few were able to or comfortable using Malmstrom vacuum extractors. Budget limitations, often make healthcare facilities in LMIC reluctant to replace equipment after a single use, resulting in staff resorting to sterilising the Kiwis using cold water, chemical sterilisation and re-using them as was found in our study and [24]. This however, eventually resulted in them failing to develop an adequate vacuum, making them unusable and also potentially risking patient to patient, cross-contamination if not properly decontaminated and sterilised. Nolens et al [24] found that following the development of a rigorous infection control procedure, they were safely able to re-use the Kiwi devices, but this was dependent on the availability of Cidex and only resulted in the devices functioning for 3-5 deployments. A solution to this problem would be the provision of a portable, hand-held device, designed to be sterilised between uses and distributed at a price that is affordable to resource-limited health systems.

##### Knowledge, skills and training

The lack of performance of AVB was in several cases attributed to a lack of knowledge and skills on the part of the clinicians. This was related to a lack of training not just in AVB but also in the basic care and assessment of women in labour, although most of the interviewees had received basic AVB training as part of the EmOC programme.

A systematic review of the effectiveness of short, competency-based EmOC training, including AVB, for healthcare providers in high, middle and low-income countries [25] found significant improvements in knowledge, skills and clinical practice following the training. The review also found however that EmOC knowledge and skills declined over time, following the training. This was corroborated by the findings of the EmOC knowledge and skills retention study carried out by Ameh et al [26] in six sub-Saharan African countries including Kenya, which showed a significant variation in the baseline knowledge and skills of maternity care providers by cadre between countries. This may suggest weakness in the pre-service training programme or/and suboptimal or lack of effective continuous medical education programmes. The Ameh et al study [26] also showed that frequent training/drills helped to maintain knowledge and skills above pre-training levels for up to 12 months and recommended periodic refresher skills training to ensure knowledge and skills are retained.

In this study, time lapse between training and practicing the skills of AVB, or extended periods of not working in maternity labour ward, were likely to result in reduced confidence levels. Although theoretically there may have been a high proportion of staff within a facility who had received training in performing AVB, the lack of practice and confidence may have been a significant barrier, or they may have been working in a non-maternity setting. Any lack of pre-service training on AVB, particularly for nurse-midwives and junior doctors cannot be adequately overcome by short, post-registration EmONC courses, particularly where there is inadequate supported clinical practice once qualified, and no mandatory, practical refresher training.

Within this study there also seemed to be a preference for external agencies to provide essential refresher training rather than senior staff within facilities taking a lead in this process, particularly the practical, skills-based aspects. This may have been for a number of reasons including lack of confidence or experience in senior member of staff who might also be reluctant to admit this. The prospect of receiving per diems (financial payments for taking part in externally funded training programmes) and an extended time away from the normal workplace, might all potentially have a bearing on it [27]. The lack of sustainability of a model using primarily external training providers, highlights the need for periodic, practical training and mentorship to be championed and provided at a local level, but for this to succeed, confident, competent mentors, and allocation of staff time to attend, are needed, as well as a method of recognising higher levels of competence.

##### Staff rotation

The rotation of staff to different departments was commented on in several of the interviews in this study and is likely to contribute to difficulties for staff in consolidating newly acquired knowledge and skills, such as AVB. Further evidence on the rotation of healthcare staff was found by Shikuku et al [28] in their review of staff retention following an EmOC training programme, in five counties in Kenya. They found that only one third of the 621 staff trained in EmOC were still working in a maternity department.

Although there is little research evidence on the effect of staff rotation on the provision of care quality, the rotation of trained and experienced staff was found to be problematic in relation to a hospital-based intervention to improve paediatric care including a 5-day training course [29] in district hospitals in Kenya. They describe, over an 18-month period, two hospitals changing their medical superintendent, one hospital changing senior paediatric nursing staff four times, and in excess of 20 paediatric nursing staff moving to other departments across three facilities. This resulted in staff experienced in the intervention package being replace with staff who had no knowledge of it, and they conclude that this rotation of staff was “the most likely general threat to the long-term success of any intervention”[29].

While these rotations may be necessary to manage the limited work force, a system is needed at hospital or district level, to ensure that those deployed to maternity wards are trained and competent to provide EmOC. Monitoring the training of healthcare providers, and the placement of EmOC trained staff to maternity wards, will help to always ensure that a critical mass of competent staff are available. This can be supplemented using a functioning and fully sustained mentorship system to promote the ongoing support and practical supervision of junior or less experienced healthcare providers. The use of mentorship can also underpin the dissemination of EmOC knowledge and skills to non-EmOC trained staff, maximising the benefits of training for women, staff, and the healthcare system.

## Conclusions

An increase in the appropriate use of AVB could avoid unnecessary cases of SSCS and the associated mortality and short- and long-term morbidity, as well as resulting in more efficient use of scarce resources and greater job satisfaction. This study highlighted a variety of issues relating to the under-utilisation of AVB in emergency obstetric care facilities in Kenya. Challenges included the lack of functional equipment, and lack of confident, competent staff on duty in labour wards within facilities. Other factors included a lack of pre-registration training on the use of AVB, together with inadequate refresher training on practical skills for AVB, for some staff, and the lack of practical leadership in some instances. The complexity of the situation means that there was no single, simple solution to the under-performance of AVB. Interventions to address the problem will need to be comprehensive and sustainable. They will need to address personal, facility-level, local community and national-level barriers, and promote a culture of AVB within facilities. It is anticipated that a prospective intervention study, that addresses health system challenges, and incorporating a package of evidence-based interventions, will improve the quality of information prevalence and outcomes of CS and AVB, as well as reducing the risk of adverse outcomes from unnecessary procedures.

## Data Availability

All data relevant to the study are included in the article.

## Acknowledgements

The authors gratefully acknowledge the members of the LSTM Kenya and UK teams for their support with this study and all of the healthcare providers who contributed their time.

## Notes

### Competing Interest Statement

Charles Ameh is an editor for Plos Global Public Health.

### Funding Statement

The study was partly funded under a grant from the UK FCDO (Reducing Maternal & Neonatal Death in Kenya, number 202549) (Charles Ameh), and partly from an award from LSTM (Fiona Dickinson). The funders had no role in study design, data collection and analysis, decision to publish, or preparation of the manuscript.

### Author Declarations

The study was approved by: LSTM REC: (21-041) Moi University IREC: (IREC/2021/115)

